# Predictors of good contraception attitude and practice among female students of television studies in Nigeria: a secondary analysis

**DOI:** 10.1101/2024.02.26.24303367

**Authors:** Philip Adewale Adeoye, Tolu Adeniji, Hadizah Abigail Agbo

## Abstract

**Background:** Globally contraceptive uptake among young people is increasing; so too is the high unmet need for family planning among this population, especially in sub-Saharan Africa. This study assessed attitude and practice item predictors of contraception among female students of Television studies in Nigeria.

**Methods:** This is a secondary analysis of a cross-sectional study among 227 female TV undergraduates in Nigeria; selected through a simple random technique by balloting. Data was analyzed using SPSS V.25 and qualitative data were presented as frequencies and proportions. Predictors of good attitude and practice were determined by multivariable logistic regression. P-value<0.05 was adjudged significant.

**Results:** most (91%) of respondents are aware of contraception. Most (94.9%) of the respondents have had unplanned pregnancies and only 42.1% had ever used contraception. Predictors of good attitude include good knowledge that female sterilization is one way to avoid pregnancy (aOR: 2.244 [95%CI: 1.170, 4.304]; p=0.015); and opportunity for switching in case of side-effect (aOR: 2.310 [95%CI: 1.166, 4.578]; p=0.016); and using both condoms and pills is very effective (aOR: 1.965 [95%CI: 1.005, 3.840]; p=0.048). The predictor of good contraceptive practice is a poor perception of the adequacy of the current method being used (aOR: 3.236 [95%CI: 1.455, 7.196]; p=0.004).

**Conclusion:** **T**his study shows that most respondents are aware of contraception. However, they show poor attitudes to and practice of contraception. There is a need to consolidate comprehensive sex education in all secondary and tertiary institutions; especially in non-science disciplines to improve the attitude and practice of contraception and ensure reproductive well-being and educational development of the girl-child. There is also the need for community action through community dialogue to improve poor contraceptive attitudes among young women.

## Introduction

Almost 30 years ago since the 1994 International Conference on Population and Development, significant progress has been made in adolescent reproductive rights and health. Adolescents are more likely to marry later, delay sexual intercourse, use contraception and delay childbirth compared to 30 years ago. This progress has however been uneven within and between countries.^1^

Though sexual debt among adolescents has declined in sub-Saharan Africa, sexual activity is highest among adolescents compared to other regions; reflecting the higher rates of child marriages and early sexual debut within marriage in the region. Due to increased experimentation with sexual activity, sexually transmitted infections (STIs) have increased among young people; with Africa having the highest prevalence for genital herpes, and gonorrhoea-with females having a higher prevalence.^1^

The proportion of young women who were married before 18 years decreased in the last 30 years; with the highest declines reported in Northern Africa, Western and Southern Asia. Also, early childbearing has declined since the early 1990s. However, this remained highest in sub-Saharan Africa Latin America and the Caribbean.^1^ Shorter interval pregnancies among young mothers pose further risks for both the mother and child.^2^

However, there has been a doubling of contraceptive use among adolescents compared to a decade ago-which currently stands at 21%. Latin America has the highest contraceptive use and a rapid rise in sub-Saharan Africa (from 4 to 15%) in the last 20 years. Contraception among unmarried sexually active adolescent girls is higher compared to their married counterparts.^2^ Condoms account for 70% of the total modern contraceptive uptake among older sexually active adolescents; while pills and injectables account for a similar 70% of modern contraceptive use among married older adolescents.^3^ Contraception is a way by which safe sex is ensured, unplanned pregnancies, early childbearing, STI, and maternal health risks are reduced.

With reduced age at menarche and onset of sexual activity, there is a heightened risk of unplanned sexual intercourse, unplanned pregnancy, unsafe abortion, severe puerperal illness, infertility maternal mortality, and adverse birth outcomes. Individual factors such as risk perception, fear of side-effects, opposition of partners, health system limitations, insufficient/lack of knowledge, sociocultural and religious factors are some of the barriers to contraception among female university undergraduates in sub-Saharan Africa.^4-8^

In Nigeria, contraceptive use increased from 6.0% in 19990 to 16.6% in 2018; with the south-west having the highest prevalence of 35.1% and the north-west with the lowest prevalence (6.7%). The north-central has a prevalence of 16.2%. In the north-central, plateau state has the highest prevalence (21.4%) of modern contraceptive use. contraceptive prevalence among sexually active unmarried women is 37%, and 28% for modern contraceptive use. Condom remains the commonest among unmarried sexually active women. There is a higher demand and unmet need for contraception among sexually unmarried compared to married women in Nigeria.^4^

However, appropriate knowledge and attitude should drive healthful and healthy reproductive behaviour across populations. Among university female undergraduates who are older young persons, the situation is the same; and in this population, knowledge, attitude and practice on contraception is at best average.^9^ It is during this period that adolescents and young people begin to experiment with sexual activities while lacking requisite knowledge on reproductive health.^10,11^ This lack of knowledge on reproductive health issues, limited implementation of comprehensive sex education in schools and poor parental knowledge has hindered discussion on sexual health issues. This makes adolescents, who are coming from a restrictive high school environment to a liberal university environment, to be vulnerable to harmful sexual behaviours.

Though there have been many studies assessing knowledge, attitude and practice among female undergraduates in Nigeria, there has not been any such study-to the best of the knowledge of the researchers-specifically among female TV studies undergraduates in Nigeria; as this population may be involved in mass media health education and risk communication for community mobilization in the future. Also, a secondary item analysis might provide insight into the levels of knowledge, attitude and practice on contraception earlier reported in a previous study.^9^

## Methods

### Study design

this is a secondary analysis of a previously published cross-sectional study;^9^ which revealed the levels of good knowledge, attittude and practice of contaception to be about average; with majority obtaining information on contraception from friends and the internet; and the commonly used contraception being condom and oral contraceptive pills. Further item analysis is to maximize the value and to provide insight into the items that were of value in predicting good attitudes and practice of contraception among the study population.

### Study area

Nigeria is a sub-Saharan African country in the western region. It is the most populous black nation on earth with an estimated population of 200million. There are about 64 million youths in Nigeria-about a third of the Nigerian population; with 51.6% female population.^12^

Plateau State, located in the North-central region of Nigeria, covers a land area of 26,899 square kilometres with an estimated population of almost 5 million. It is a multi-ethnic state; with English and Hausa being the commonly spoken languages. Because of its high altitude, it has a near-temperate climate. It has 5 specialized higher educational institutions out of the 12 higher institutions awarding post-secondary school certificates.^13^

NTA TV College is a specialized higher educational institution established in 1980 to address human resource challenges in the TV/broadcast industry. It is located in Rayfield, Jos-metropolis, Plateau State. It is the only TV College offering diplomas and degrees in TV journalism and production in Nigeria. It is currently an affiliate of Ahmadu Bello University, Zaria, Nigeria.^14^

### Study population

these are female NTA TV College students who consented to participate in this study. The primary study shows that this study participants have an average age of 21.9 (± 2.64) years; with most (91.2%) not living with a spouse or partner; Christian (91.2%); almost two-thirds (58.1%) under the degree programme; and half (51%) with a monthly income below the minimum wage; almost two-thirds are resident of the state and a majority (76.7%) living off-campus when in school and 69.6% in the TV journalism course.^9^

### Sample size determination

sample size was determined using Cochran’s sample size formula: n=z^2^pq/d^2^;^15^ where n=minimum sample size; z=standard normal deviate at 95% confidence interval=1.96; p=proportion of female undergraduates with good knowledge of contraception=0.84;^16^ q=alternate probability =1-0.84=0.16; d= precision at 5% error=0.05. Therefore, the minimum sample size is 206. Correcting for 10% nonresponse makes the total sample size=227.

### Sampling technique

the female students were selected using a simple random sampling technique by balloting for all six levels during classes in the college using the proportion-to-size approach.

### Study tool/data collection method

data collection was by the use of semi-structured self-administered questionnaires following informed written consent by each selected participant. The questionnaire has four sections: knowledge, attitude and practice of contraception and sexual behaviour. It was piloted and pretested in a comparable institution (National Film Institute) before the commencement of the study to detect errors, test for fidelity and feasibility and participant understanding of the items of the questionnaire.

### Data analysis

data was entered in and analyzed by SPSS V.25 (IBM, Armonk, NY, USA). Qualitative variables were presented as frequencies and proportions. Simple logistic regression was used to determine factors that are associated with good attitude and practice to contraception; and to determine candidate variables for the multivariable analysis. Variables with less than 10% probability of error were selected and added into the multivariable omnibus model to determine the predictors of good attitude and practice of contraception. P-value<0.05 was adjudged statistically significant.

### Ethics approval and consent to participate

ethics approval was obtained from the JUTH Research and Ethics Committee (JUTHH/DCS/IREC/127/XXXI/2619). Written informed consent was obtained from each of the study participants before the commencement of the study; which was voluntary and withdrawal was allowed at any stage of the research process. There is no potential hazard to study participants.

## Results

***Table 1*** above shows that the majority have heard about family planning; and know that health education is important for its use. About two-thirds know that pills do not guarantee 100% protection; and know that using both the condom and pills is very effective. Below half of the respondents know that pills may increase the risk of breast and cervical cancers; know the common side-effects of pills and know that female sterilization is a method of contraception. About one-third know that condoms do prevent STIs; increase the risk of breast cancer among those using oral contraceptive pills; Depo given every 3 months and a switch might be necessary if there is a side-effect. Only 28% know that pills become ineffective with 2-3 omissions or missed intake.

**Table 1:** Knowledge of Contraception among female students at NTA TV College Jos.

| Variable | Frequency (%) |
| --- | --- |
| **Ever heard of contraception (n=214)** |  |
| Yes (good knowledge) | 195 (91.1) |
| No/ I don't know (poor knowledge) | 19 (8.9) |
| **Pills are effective with 2-3 omissions (n=211)** |  |
| No (good knowledge) | 59 (28.0) |
| Yes/I don't know (poor knowledge) | 152 (72.0) |
| **Female sterilization is one way to avoid pregnancy (n=213)** |  |
| Yes (good knowledge) | 98 (46.0) |
| No/ I don't know (poor knowledge) | 115 (54.0) |
| **Health education is important for its use (n=215)** |  |
| Yes (good knowledge) | 191 (88.8) |
| No/ I don't know (poor knowledge) | 24 (11.2) |
| **Pills guarantee 100% protection (n=214)** |  |
| No (good knowledge) | 133 (62.1) |
| Yes/I don't know (poor knowledge) | 81 (37.9) |
| **Condoms do not prevent STI (n=214)** |  |
| No (good knowledge) | 78 (36.4) |
| Yes/I don't know (poor knowledge) | 136 (63.6) |
| **Pill common side-effect include mood swings and weight gain (n=213)** |  |
| Yes (good knowledge) | 104 (48.8) |
| No/ I don't know (poor knowledge) | 109 (51.2) |
| **Increased risk of breast cancer with those taking estrogen-containing contraceptives (n=217)** |  |
| Yes (good knowledge) | 78 (35.9) |
| No/ I don't know (poor knowledge) | 139 (64.1) |
| **Those using depo must get injections every 3 months (n=213)** |  |
| Yes (good knowledge) | 70 (32.9) |
| No/ I don't know (poor knowledge) | 143 (67.1) |
| **If there is a side-effect; switching might help (n=217)** |  |
| Yes (good knowledge) | 70 (32.3) |
| No/ I don't know (poor knowledge) | 147 (67.7) |
| **Using both condoms and pills is very effective (n=216)** |  |
| Yes (good knowledge) | 124 (57.4) |
| No/ I don't know (poor knowledge) | 92 (42.6) |
| **Pills increase the risk of cervical cancer (n=217)** |  |
| Yes (good knowledge) | 100 (46.1) |
| No/ I don't know (poor knowledge) | 117 (53.9) |

***Table 2*** above shows that the majority, or most, of the tertiary students have poor attitudes to issues regarding contraception. For example, only a minority of respondents believe that contraception provides a sense of safety; important in child spacing and child health; and the perception of the adequacy of the contraception in use.

**Table 2:** Attitude to contraception among female students at NTA TV College Jos.

| Variables | Frequency<br>(percentage) |
| --- | --- |
| **It should be used to limit my number of children (n=214)** |  |
| Poor attitude (Neutral/Disagree/Strongly Disagree) | 176 (82.2) |
| Good attitude (Strongly Agree/Agree) | 38 (17.8) |
| **Should be used to increase the time interval between my child's births (n=214)** |  |
| Poor attitude (Neutral/Disagree/Strongly Disagree) | 179 (83.6) |
| Good attitude (Strongly Agree/Agree) | 35 (16.4) |
| **Spacing will allow a child to be healthier (n=216)** |  |
| Poor attitude (Neutral/Disagree/Strongly Disagree) | 190 (88.0) |
| Good attitude (Strongly Agree/Agree) | 26 (12.0) |
| **Ideal age for having a first child is between 20 and 30 years of age (n=211)** |  |
| Poor attitude (Neutral/Disagree/Strongly Disagree) | 180 (85.3) |
| Good attitude (Strongly Agree/Agree) | 31 (14.7) |
| **Ideal number of children between 3 and 5 (n=212)** |  |
| Poor attitude (Neutral/Disagree/Strongly Disagree) | 181 (85.4) |
| Good attitude (Strongly Agree/Agree) | 31 (14.6) |
| **Contraceptives provide a sense of safety (n=211)** |  |
| Poor attitude (Neutral/Disagree/Strongly Disagree) | 176 (83.4) |
| Good attitude (Strongly Agree/Agree) | 35 (16.6) |
| **The method I am using is adequate (n=203)** |  |
| Poor attitude (Neutral/Disagree/Strongly Disagree) | 149 (73.4) |
| Good attitude (Strongly Agree/Agree) | 54 (26.6) |
| **Benefits males too (n=209)** |  |
| Poor attitude (Neutral/Disagree/Strongly Disagree) | 150 (71.8) |
| Good attitude (Strongly Agree/Agree) | 59 (28.2) |
| **Discussion about contraception with partner is embarrassing (n=209)** |  |
| Poor (Strongly Agree/Agree/Neutral) | 165 (78.9) |
| Good (Disagree/Strongly Disagree) | 44 (21.1) |
| **Partner does not approve of my use of contraceptives (n=206)** |  |
| Poor (Strongly Agree/Agree/Neutral) | 159(77.2) |
| Good (Disagree/Strongly Disagree) | 47 (22.8) |
| **Can protect the health of family and community (n=212)** |  |
| Poor attitude (Neutral/Disagree/Strongly Disagree) | 173 (81.6) |
| Good attitude (Strongly Agree/Agree) | 39 (18.4) |
| **Religious beliefs can prevent women from using contraceptives (n=213)** |  |
| Poor attitude (Neutral/Disagree/Strongly Disagree) | 169 (79.3) |
| Good attitude (Strongly Agree/Agree) | 44 (20.7) |
| **Cultural beliefs can prevent women from using contraceptives (n=211)** |  |
| Poor attitude (Neutral/Disagree/Strongly Disagree) | 179 (84.8) |
| Good attitude (Strongly Agree/Agree) | 32 (15.2) |
| **Partner's objection can prevent women from using contraceptives (n=216)** |  |
| Poor attitude (Neutral/Disagree/Strongly Disagree) | 180 (83.3) |
| Good attitude (Strongly Agree/Agree) | 36 (16.7) |
| **Change in male attitude can improve contraceptive use (n=216)** |  |
| Poor attitude (Neutral/Disagree/Strongly Disagree) | 161 (74.5) |
| Good attitude (Strongly Agree/Agree) | 55 (25.5) |

***Table 3*** above shows that more than two-thirds always visit health facilities for contraception; always use different types of contraceptives per time; and always practice natural methods when not using any modern method. About half always use contraceptives to prevent unplanned pregnancy or when they do not want to get pregnant. Only about 5.1% had never had an unplanned pregnancy due to non-use, and 6.6% had never changed their current contraceptive from time to time.

**Table 3:**
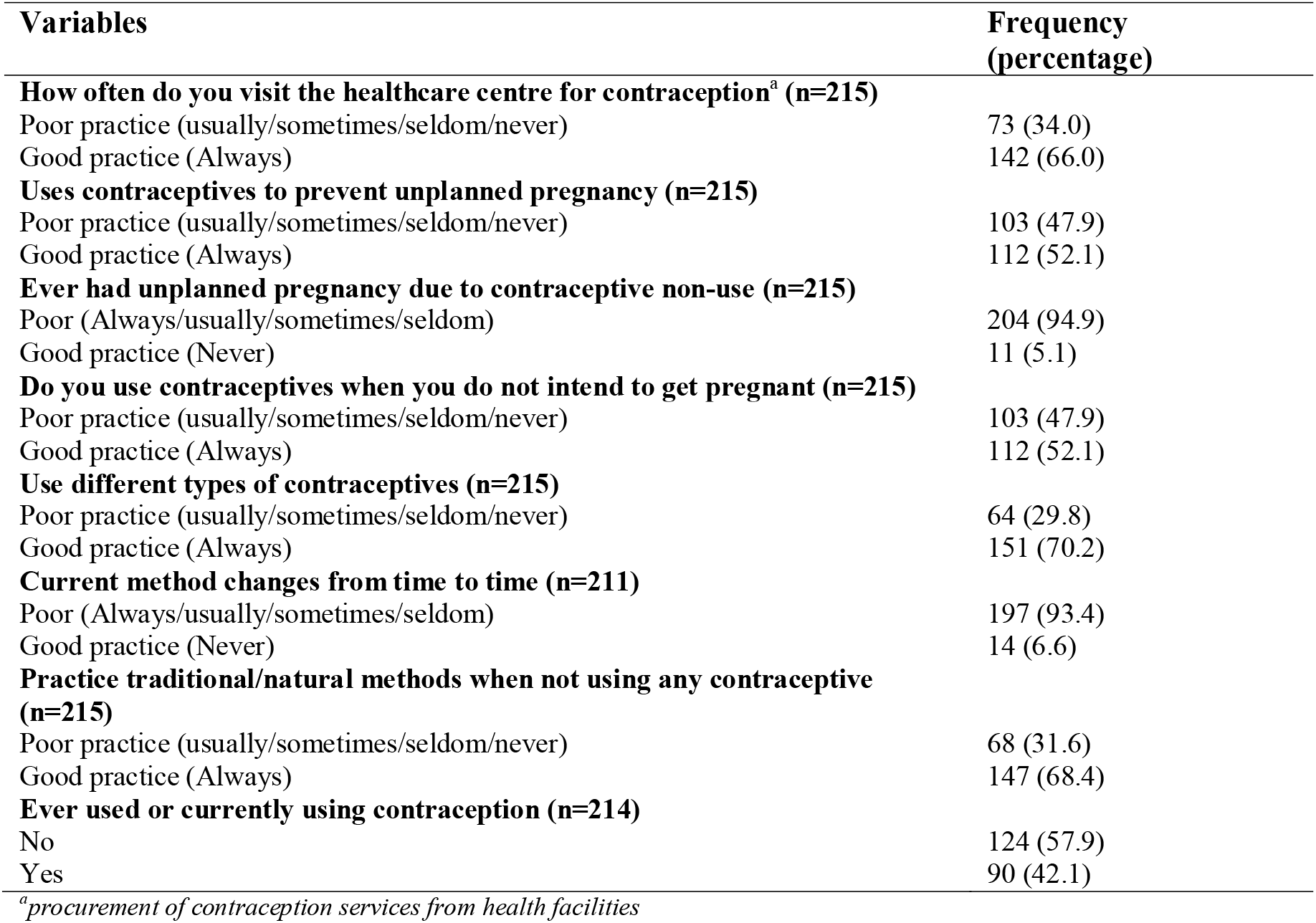
Practice of contraception among female students at NTA TV College Jos.

***Table 4*** below shows individuals who know that female sterilization is a method of contraception are twice significantly more likely to have a good attitude towards contraception compared to those who do not know. Those who know that switching to another contraceptive might help with dealing with the side-effects of a method are twice significantly more likely to have a good attitude towards contraception compared to those who do not know. Those who know that using both condoms and pills is very effective are almost twice significantly more likely to have a good attitude towards contraception compared to those who do not know.

**Table 4:**
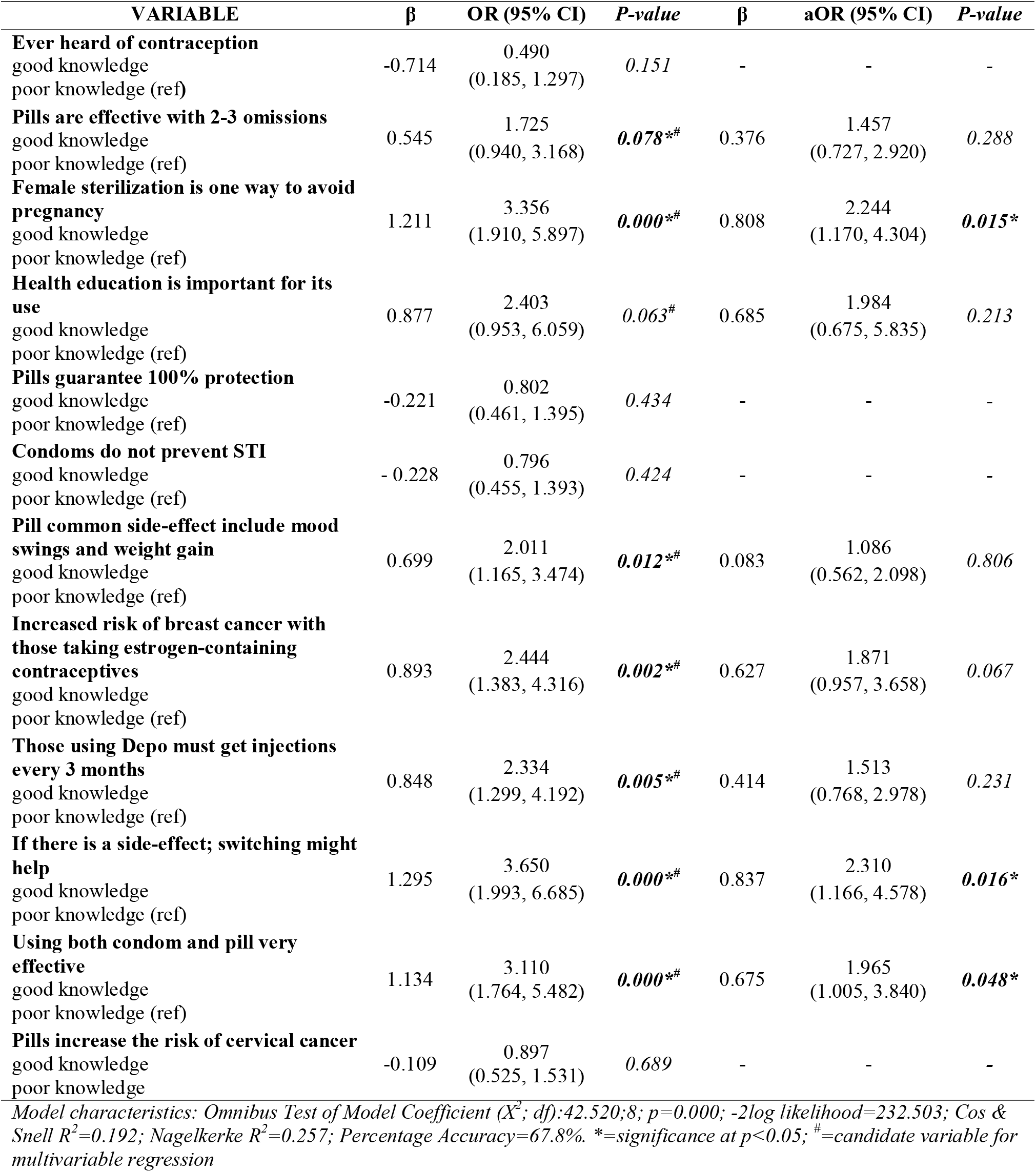
knowledge predictors of good attitude to contraception among female students at NTA TV College Jos.

| VARIABLE | $\beta$ | OR (95% CI) | P-value | $\beta$ | aOR (95% CI) | P-value |
| --- | --- | --- | --- | --- | --- | --- |
| **Ever heard of contraception** |  |  |  |  |  |  |
| good knowledge | -0.714 | 0.490<br>(0.185, 1.297) | 0.151 | - | - | - |
| poor knowledge (ref) |  |  |  |  |  |  |
| **Pills are effective with 2-3 omissions** |  |  |  |  |  |  |
| good knowledge | 0.545 | 1.725<br>(0.940, 3.168) | 0.078*<sup>#</sup> | 0.376 | 1.457<br>(0.727, 2.920) | 0.288 |
| poor knowledge (ref) |  |  |  |  |  |  |
| **Female sterilization is one way to avoid pregnancy** |  |  |  |  |  |  |
| good knowledge | 1.211 | 3.356<br>(1.910, 5.897) | 0.000*<sup>#</sup> | 0.808 | 2.244<br>(1.170, 4.304) | 0.015* |
| poor knowledge (ref) |  |  |  |  |  |  |
| **Health education is important for its use** |  |  |  |  |  |  |
| good knowledge | 0.877 | 2.403<br>(0.953, 6.059) | 0.063<sup>#</sup> | 0.685 | 1.984<br>(0.675, 5.835) | 0.213 |
| poor knowledge (ref) |  |  |  |  |  |  |
| **Pills guarantee 100% protection** |  |  |  |  |  |  |
| good knowledge | -0.221 | 0.802<br>(0.461, 1.395) | 0.434 | - | - | - |
| poor knowledge (ref) |  |  |  |  |  |  |
| **Condoms do not prevent STI** |  |  |  |  |  |  |
| good knowledge | -0.228 | 0.796<br>(0.455, 1.393) | 0.424 | - | - | - |
| poor knowledge (ref) |  |  |  |  |  |  |
| **Pill common side-effect include mood swings and weight gain** |  |  |  |  |  |  |
| good knowledge | 0.699 | 2.011<br>(1.165, 3.474) | 0.012*<sup>#</sup> | 0.083 | 1.086<br>(0.562, 2.098) | 0.806 |
| poor knowledge (ref) |  |  |  |  |  |  |
| **Increased risk of breast cancer with those taking estrogen-containing contraceptives** |  |  |  |  |  |  |
| good knowledge | 0.893 | 2.444<br>(1.383, 4.316) | 0.002*<sup>#</sup> | 0.627 | 1.871<br>(0.957, 3.658) | 0.067 |
| poor knowledge (ref) |  |  |  |  |  |  |
| **Those using Depo must get injections every 3 months** |  |  |  |  |  |  |
| good knowledge | 0.848 | 2.334<br>(1.299, 4.192) | 0.005*<sup>#</sup> | 0.414 | 1.513<br>(0.768, 2.978) | 0.231 |
| poor knowledge (ref) |  |  |  |  |  |  |
| **If there is a side-effect; switching might help** |  |  |  |  |  |  |
| good knowledge | 1.295 | 3.650<br>(1.993, 6.685) | 0.000*<sup>#</sup> | 0.837 | 2.310<br>(1.166, 4.578) | 0.016* |
| poor knowledge (ref) |  |  |  |  |  |  |
| **Using both condom and pill very effective** |  |  |  |  |  |  |
| good knowledge | 1.134 | 3.110<br>(1.764, 5.482) | 0.000*<sup>#</sup> | 0.675 | 1.965<br>(1.005, 3.840) | 0.048* |
| poor knowledge (ref) |  |  |  |  |  |  |
| **Pills increase the risk of cervical cancer** |  |  |  |  |  |  |
| good knowledge | -0.109 | 0.897<br>(0.525, 1.531) | 0.689 | - | - | - |
| poor knowledge |  |  |  |  |  |  |

Model characteristics: Omnibus Test of Model Coefficient ( $X^2$ ; df):42.520;8; $p=0.000$ ; -2log likelihood=232.503; Cos & Snell $R^2=0.192$ ; Nagelkerke $R^2=0.257$ ; Percentage Accuracy=67.8%. \*=significance at $p<0.05$ ; <sup>#</sup>=candidate variable for multivariable regression

**Table 5** below shows that those who have poor attitudes towards the adequacy of their current contraception are three times significantly more likely to have good contraceptive practices compared to those who have good attitudes.

**Table 5:** knowledge and attitude predictors of good contraception practice among female students at NTA TV College Jos.

| VARIABLE | B | OR (90% CI) | P-value | β | aOR (95% CI) | P-value |
| --- | --- | --- | --- | --- | --- | --- |
| **Ever heard of contraception** |  |  |  |  |  |  |
| good knowledge | -0.075 | 0.928<br>(0.361, 2.384) | 0.877 | - | - | - |
| poor knowledge (ref) |  |  |  |  |  |  |
| **Pills are effective with 2-3 omissions** |  |  |  |  |  |  |
| good knowledge | -0.302 | 0.740<br>(0.405, 1.352) | 0.327 | - | - | - |
| poor knowledge (ref) |  |  |  |  |  |  |
| **Female sterilization is one way to avoid pregnancy** |  |  |  |  |  |  |
| good knowledge | 0.175 | 1.191<br>(0.694, 2.043) | 0.525 | - | - | - |
| poor knowledge (ref) |  |  |  |  |  |  |
| **Health education is important for its use** |  |  |  |  |  |  |
| good knowledge | 0.031 | 1.032<br>(0.442, 2.412) | 0.942 | - | - | - |
| poor knowledge (ref) |  |  |  |  |  |  |
| **Pills guarantee 100% protection** |  |  |  |  |  |  |
| good knowledge | -0.329 | 0.720<br>(0.413, 1.254) | 0.246 | - | - | - |
| poor knowledge (ref) |  |  |  |  |  |  |
| **Condoms do not prevent STI** |  |  |  |  |  |  |
| good knowledge | 0.570 | 1.767<br>(1.006, 3.106) | **0.048\*<sup>#</sup>** | 0.532 | 1.702<br>(0.885, 3.275) | 0.111 |
| poor knowledge (ref) |  |  |  |  |  |  |
| **Pill common side-effect include mood swings and weight gain** |  |  |  |  |  |  |
| good knowledge | 0.625 | 1.868<br>(1.084, 3.217) | **0.024\*<sup>#</sup>** | 0.419 | 1.520<br>(0.778, 2.970) | 0.221 |
| poor knowledge (ref) |  |  |  |  |  |  |
| **Increased risk of breast cancer with those taking estrogen-containing contraceptives** |  |  |  |  |  |  |
| good knowledge | 0.604 | 1.829<br>(1.041, 3.212) | **0.036\*<sup>#</sup>** | 0.268 | 1.307<br>(0.654, 2.611) | 0.448 |
| poor knowledge (ref) |  |  |  |  |  |  |
| **Those using Depo must get injections every 3 months** |  |  |  |  |  |  |
| good knowledge | 0.532 | 1.701<br>(0.953, 3.039) | 0.073<sup>#</sup> | 0.141 | 1.152<br>(0.580, 2.287) | 0.687 |
| poor knowledge (ref) |  |  |  |  |  |  |
| **If there is a side-effect; switching might help** |  |  |  |  |  |  |
| good knowledge | 0.731 | 2.077<br>(1.159, 3.723) | **0.014\*<sup>#</sup>** | 0.287 | 1.333<br>(0.670, 2.654) | 0.413 |
| poor knowledge (ref) |  |  |  |  |  |  |
| **Using both condoms and pills is very effective** |  |  |  |  |  |  |
| good knowledge | 0.880 | 2.410<br>(1.386, 4.190) | **0.002\*<sup>#</sup>** | 0.501 | 1.650<br>(0.849, 3.206) | 0.140 |
| poor knowledge (ref) |  |  |  |  |  |  |
| **Pills increase the risk of cervical cancer** |  |  |  |  |  |  |
| good knowledge | 0.246 | 1.279<br>(0.749, 2.183) | 0.368 | - | - | - |
| poor knowledge (ref) |  |  |  |  |  |  |
| **It should be used to limit the number of children** |  |  |  |  |  |  |
| poor attitude | 0.151 | 1.163<br>(0.576, 2.346) | 0.674 | - | - | - |
| good attitude (ref) |  |  |  |  |  |  |

**Table 5 (contd): knowledge and attitude predictors of good contraception practice among female students** **at NTA TV College Jos**
| VARIABLE | B | OR (90% CI) | P-value | $\beta$ | aOR (95% CI) | P-value |
| --- | --- | --- | --- | --- | --- | --- |
| **Should be used to increase the time interval between my child's births** |  |  |  |  |  |  |
| poor attitude | 0.250 | 1.284<br>(0.621, 2.656) | 0.500 | - | - | - |
| good attitude (ref) |  |  |  |  |  |  |
| **Spacing will allow a child to be healthier** |  |  |  |  |  |  |
| poor attitude | 0.217 | 1.243<br>(0.546, 2.827) | 0.604 | - | - | - |
| good attitude (ref) |  |  |  |  |  |  |
| **Ideal age for having a first child is between 20 and 30 years of age** |  |  |  |  |  |  |
| poor attitude | 0.239 | 1.269<br>(0.591, 2.729) | 0.541 | - | - | - |
| good attitude (ref) |  |  |  |  |  |  |
| **Ideal number of children between 3 and 5** |  |  |  |  |  |  |
| poor attitude | 0.060 | 2.886<br>(1.260, 6.611) | 0.012*<sup>#</sup> | 0.427 | 1.532<br>(0.555, 4.231) | 0.411 |
| good attitude (ref) |  |  |  |  |  |  |
| **Contraceptives provide a sense of safety** |  |  |  |  |  |  |
| poor attitude | 0.401 | 1.494<br>(0.719, 3.106) | 0.282 | - | - | - |
| good attitude (ref) |  |  |  |  |  |  |
| **The method I am using is adequate** |  |  |  |  |  |  |
| poor attitude | 1.089 | 2.971<br>(1.536, 5.748) | 0.001*<sup>#</sup> | 1.174 | 3.236<br>(1.455, 7.196) | 0.004* |
| good attitude (ref) |  |  |  |  |  |  |
| **Benefits males too** |  |  |  |  |  |  |
| poor attitude | -0.388 | 0.679<br>(0.370, 1.246) | 0.211 | - | - | - |
| good attitude (ref) |  |  |  |  |  |  |
| **Discussion about contraception with partner is embarrassing** |  |  |  |  |  |  |
| poor attitude | -0.219 | 0.804<br>(0.412, 1.566) | 0.521 | - | - | - |
| good attitude (ref) |  |  |  |  |  |  |
| **Partner does not approve of my use of contraceptives** |  |  |  |  |  |  |
| poor attitude | 0.125 | 1.134<br>(0.590, 2.180) | 0.707 | - | - | - |
| good attitude (ref) |  |  |  |  |  |  |
| **Can protect the health of family and community** |  |  |  |  |  |  |
| poor attitude | 0.490 | 1.633<br>(0.807, 3.303) | 0.173 | - | - | - |
| good attitude (ref) |  |  |  |  |  |  |
| **Religious beliefs can prevent women from using contraceptives** |  |  |  |  |  |  |
| poor attitude | -0.427 | 0.653<br>(0.333, 1.278) | 0.213 | - | - | - |
| good attitude (ref) |  |  |  |  |  |  |
| **Cultural beliefs can prevent women from using contraceptives** |  |  |  |  |  |  |
| poor attitude | -0.544 | 0.580<br>(0.268, 1.257) | 0.168 | - | - | - |
| good attitude (ref) |  |  |  |  |  |  |
| **Partner's objection can prevent women from using contraceptives** |  |  |  |  |  |  |
| poor attitude | -0.637 | 0.529<br>(0.252, 1.109) | 0.092<sup>#</sup> | -0.800 | 0.449<br>(0.183, 1.102) | 0.080 |
| good attitude (ref) |  |  |  |  |  |  |
| **Change in male attitude can improve contraceptive use** |  |  |  |  |  |  |
| poor attitude | -0.594 | 0.552<br>(0.295, 1.033) | 0.063<sup>#</sup> | -0.722 | 0.486<br>(0.221, 1.069) | 0.073 |
| good attitude (ref) |  |  |  |  |  |  |

Model characteristics: Omnibus Test of Model Coefficient ( $X^2$ ; df):33.063;10; $p=0.000$ ; -2log likelihood=227.560; Cos & Snell $R^2=0.161$ ; Nagelkerke $R^2=0.215$ ; Hosmer-Lemeshow $R^2 p=0.228$ ; percentage accuracy=68.1%. \*=significance at $p<0.05$ ; <sup>#</sup>=candidate variable for multivariable regression

## Discussion

Most of the respondents, in this study, had heard of contraception. This is similar to those observed among similar populations in Dodoma-Tanzania, states in North-Central Nigeria, Thohoyandou-South Africa, and Suva-Fiji.^5,6,17-19^ This may be due to increased sensitization and community mobilization on contraception in most countries of the world; especially in lower- and middle-income countries (LMICs).^20^ This will need to be sustained especially in hard-to-reach regions of the world.

Furthermore, almost half of the respondents know that female sterilization is an option to avoid pregnancy. Lower values and similarly average proportions have been reported similar proportion in Dodoma-Rwanda and Nigeria among similar populations.^4,5^ This variation may be due to limited practical in-depth details about contraception; especially at the grassroots.

Health education has been reported by the majority of these study respondents as important for successful contraceptive use. This is similar to those observed in Kigali-Rwanda, Gaborone-Botswana and Suva-Fiji among similar populations.^6,21,22^ This shows a corresponding enthusiasm that a very high awareness might have created.

Below average of respondents know that risks associated with OCP include breast, and cervical cancers. A similar observation was reported among similar populations in Suva-Fiji.^6^ However, higher proportions were reported in Thohoyandou-South Africa.^18^ This may be due to improved reporting and access to some cancer screening services in the Southern Africa region in Sub-Saharan Africa.^23^

About a third of respondents know that switching may help if there is a side-effect; a higher proportion of knowledge was reported among women in Suva-Fiji.^6^ Despite this, only a small proportion of contraceptive users switch among Nigerian unmarried sexually active women in a recent survey.^4^ This might be due to improved selection of appropriate contraceptives for women of reproductive age group which had probably minimized the side-effects associated with contraceptives.

Most respondents of this study have poor attitude towards contraception across contraceptive attitude. This is similar to the attitude of undergraduates in Pa Nang-Vietnam and Malaysia.^24,25^ This may be as are result of socio-cultural factors that still surround contraception in many LMiCs.^20^

The majority of respondents do not believe that spacing will make their children healthy; and found it unnecessary to increase birth intervals. Contrary opinion was expressed by an equal proportion of women in Suva -Fiji.^6^ A similar opinion in support of definite birth spacing was reported in Senegal, where *Nef* (short birth interval) is frowned upon and stigmatizing; and in line with evidence of adverse birth outcomes for the mother and child.^26^ Thus, there is a need for culture-specific tailoring of contraceptive education and mobilization; as not all caps fit all.

Almost three-quarters do not believe that contraceptives benefit males too. This is contrary to that which is observed in similar populations in Kigali-Rwanda and Suva-Fiji;^6,22^ where more than three-quarters have good attitude that males benefit also from contraception. This may explain why contraception is thought to be in the main interest of the woman and some Nigerian men will be involved in family planning programmes to show love and support for their wives.^7^

Majority of the female undergraduates in the study site disagree that the ideal age for the onset of childbearing should be between 20 and 30 years of age. This is, however, different for the majority of women in Suva-Fiji.^6^ This might be due to the widespread teenage pregnancies; which are being normalized in many states in Northern Nigeria childhood and teenage pregnancies and marriage also have the backing of cultural and religious factors in many communities in Nigeria. The increasing numbers of out-of-school teenagers in Nigeria might have also contributed to the normalizing of teenage pregnancies and marriages in Nigeria.^28^

From this study, a majority do not believe that their current contraceptive is adequate. This is contrary to the perception expressed by a similar population in Suva-Fiji.^6^ This might be responsible for the use of different types and frequent changes of contraceptives from time to time among the study population which was not seen among similar populations in Suva-Fiji. Therefore, there may be a need for increased confidence in contraception via community mobilization and participation in contraception activities.

This study shows that many of the study participants visit the healthcare centres for contraceptive uptake. This might be due to the freely available contraceptives at these public healthcare facilities in the state in which the study was done.^29^ This is however contrary to those by contrary to those reported by similar populations in Dodoma-Tanzania, Suva-Fiji and women of reproductive age in many LMiCs.^4,5,7^ This might also be due to cultural and social factors surrounding procurement of contraceptives by young people from the public sector which might be ebbing in places where contraceptives are procured from public places. Procuring contraceptives by this population from sites outside the public sector may affect accurate reporting of contraceptive use as these sites are not routinely captured in national data.^30^

Most of the study participants have had unplanned pregnancies due to the non-use of contraception. This is, however, contrary to a similar population in Suva-Fiji whose majority had never had unplanned pregnancy.^6^ This might be responsible for the use and intention to use. Contraception, subsequently, by more than half of the respondents. There is therefore the need to strengthen the Federal Ministry of Education’s family life and HIV/AIDS Education early in the education sector to prevent the adverse effects of teenage pregnancies.^31^ There is also the need to engage and commit families in the design and implementation of comprehensive sex education.^8^

Majority of the study participants practised traditional/natural methods (TCM) when not using any modern contraception. This is similar to those reported among similar populations in Kilimanjaro-Tanzania and Suva-Fiji.^6,32^ This is however contrary to those reported among similar populations in Kigali-Rwanda, Romania, and Kano-Nigeria.^33-35^ This may be as a result of unmet needs, safety concerns, side-effect and opposition to modern contraceptive use by partners; and the effectiveness of TCM.^35,36^

Less than half of respondents have ever used contraception among the study population. This is similar to those reported among similar populations in Kilimanjaro-Tanzania and Nigeria.^4,32^ This is however different for similar populations in Kigali-Rwanda, Romania and Makurdi-Nigeria.^19,22,33^ This may be due to social and cultural, health systems barriers that hinder young people from accessing contraceptive services in many LMiCs.^4,20^ This might have also explained the higher number of unplanned pregnancies experienced by this study population.^20,23^

This study shows that knowing that female sterilization is one way to prevent pregnancy is a prediction of good contraceptive attitude. Though studies on female sterilization are generally low;^37^ it can be hypothesized that a high knowledge of female sterilization is correlated with a high level of knowledge of other contraceptive methods because knowledge about female sterilization is lower compared to other methods in many populations. Increased knowledge of female sterilization is often associated with increased acceptance and uptake, all things being equal it has been shown compared to other methods, to improve mood and sex life.^22,37^ Generally, it has been said to be the most common family planning method globally among women of reproductive group.^38^

The knowledge that switching might help in the occurrence of side effects is a significant predictor of good contraceptive attitude. This may increase improved sense of safety; self-efficacy and perception that clients have control over their reproductive health. The need to conceive in the near future, fewer side-effects and the expensive nature of long-acting contraceptives are some of the reasons why long-acting reversible contraceptives have low patronage compared to the short-acting contraceptive methods in many LMiCs.^20^

This study also shows that more than half of the respondents know that combined use of condoms and pills is very effective; and this knowledge is a significant predictor of good attitude towards contraception. This combination maximises the contraceptive benefits of pregnancy and STI prevention among adolescents and young adults.

The doubt that the method clients are using is adequate is a significant predictor of good contraceptive use. This might be due to past unplanned pregnancies reported in this study population and frequent changes and use of different types of contraceptives among this population. This might have improved attendance at follow-ups after contraceptive uptake where information on correct use will be consolidated and prompt report of side-effects and cross-confirmation of any contraceptive information shared might have been responsible for good practice despite less confidence in the method being used. However, the health system should continue to boost self-confidence in this population to ensure sustainability of good contraceptive use.

This study cannot be generalized to the total female undergraduate population in other higher institutions since the population is from a specific discipline that may be involved in health communication and education via mass media in the future. The use of questionnaires might bias participant’s responses as sensitive questions are included in the items. However, respondents were provided a private setting, adequate preamble, use of anonymous research assistants and ethical conduct of the interview made respondents relaxed to answer the questions to the best of their abilities. Also, due to the cross-sectional nature of the study, temporality cannot be established. Our findings have been one of the few among similar populations in literature and have huge policies for health promotion; now and in the future. This is a cross-sectional study and therefore causality cannot be established

## Conclusion

This study showed that though most respondents are aware of contraception, they generally show poor attitude to contraception. Majority had a history of unplanned pregnancies and few had ever used contraception; though with a willingness to do so. There is, therefore, the need to consolidate early and appropriate family life and HIV/AIDS education in all levels of formal education to improve family planning knowledge, attitude and practice in many young people. There is also a need for community dialogue to improve the poor contraceptive attitude and the need to involve families and communities in the design and implementation of contraception activities. This will ensure reproductive health and all-round development of the girl-child.

## HIGHLIGHTS

- Poor attitude and practice of contraception, has been one of the main driven driving up unsustainable population growth in low- and middle-income countries.
- Most of the study participants have had past history of unplanned pregnancy
- Less than half have ever used contraception
- Knowledge of female sterilization, risk of breast cancer and opportunity of switching predicts good attitude
- Perception about the adequacy of current method is a significant predictor of good contraceptive practice

## Data Availability

All data produced in the present study are available upon reasonable request to the authors

## Acknowledgements

We are grateful to the students of NTA TV College of Nigeria for their willingness to participate in the study. We also grateful for the research assistants that patiently carried out the data collection.

## Contributions

Conceptualization and design: PAA, HAO; Data analysis and interpretation: PAA; Drafting and critical review: HAO, PAA, TA; Final approval: HAO, PAA, TA.

